# Comparison of the 2021 COVID-19 Roadmap Projections against Public Health Data in England

**DOI:** 10.1101/2022.03.17.22272535

**Authors:** Matt J. Keeling, Louise Dyson, Michael J. Tildesley, Edward M. Hill, Samuel Moore

## Abstract

Control and mitigation of the COVID-19 pandemic in England has relied on a combination of vaccination and non-pharmaceutical interventions (NPIs). Some of these NPIs are extremely costly (economically and socially), so it was important to relax these promptly without overwhelming already burdened health services. The eventual policy was a Roadmap of four relaxation steps throughout 2021, taking England from lock-down to the cessation of all restrictions on social interaction. In a series of six Roadmap documents generated throughout 2021, models assessed the potential risk of each relaxation step. Here we show that the model projections generated a reliable estimation of medium-term hospital admission trends, with the data points up to September 2021 generally lying within our 95% prediction intervals. The greatest uncertainties in the modelled scenarios came from vaccine efficacy estimates against novel variants, and from assumptions about human behaviour in the face of changing restrictions and risk.

## Introduction

Mathematical and statistical models have been an important tool in understanding the behaviour of the COVID-19 pandemic in England and extrapolating this understanding into the near future [1–9]. Such model projections rely on robust estimates of epidemiological parameters (usually inferred by matching models to data), estimates of vaccine characteristics (often from external sources) and assumptions about population behaviour in terms of testing, isolation, social mixing and vaccine uptake - errors in any one of these can affect the projected exponential growth rate and hence the medium and longer term dynamics. There has been considerable interest from the media and the general public about the accuracy of model projections, and whether these are generally far more pessimistic than what transpired [10]. While model projections are not forecasts – they are designed to illustrate “what if” scenarios [11] – it is still important that such projections remain broadly consistent with the realised epidemic if they are to be of use in public health planning.

Here we focus on the Warwick SARS-CoV-2 transmission model [5, 7, 13] and its use in the six Roadmap documents developed throughout 2021 [14–19] that helped to shape the relaxation of restrictions in England after the January 2021 lockdown through a series of Steps, as detailed below:

- Lockdown (4th January - 7th March 2021). Stay at home order: Shopping for basic necessities only; non-essential retail, hospitality and personal care services closed. Work from home where possible. Can leave home for exercise (with household, support bubble or one other person), to meet support bubble, or to seek medical care. Remote learning in schools except key workers and vulnerable. Clinically extremely vulnerable advised to shield.
- Step 1a (8th March 2021). Schools re-open, with twice-weekly testing of staff and pupils.
- Step 1b (29th March 2021). Meeting outdoors with 6 people or 2 households allowed. Outdoor sports facilities can re-open.
- Step 2 (12th April 2021). Non-essential retail, personal care and public buildings can re-open. Outdoor table service in hospitality venues.
- Step 3 (17th May 2021). Meeting outdoors with up to 30 people, and indoors with 6 people or 2 households allowed. Most businesses can reopen, including indoor hospitality.
- Step 4 (19th July 2021). Originally set for 21st June, after this step all legal limits on social contact were removed.

The Roadmap documents chronicle our changing understanding of SARS-CoV-2, vaccination and human behaviour over this period. Throughout we use the daily number of hospital admissions as our main metric of interest, but also consider hospital occupancy and deaths as key epidemiological quantities. We do not seek to perform a rigorous statistical comparison between model projections and data, since projections cannot be expected to account for the multitude of unknowable factors (human behaviour, advances in treatment, novel variants); the further into the future the models consider, the greater the compounded impact of multiple uncertainties due to there being more of these unknowable possibilities. In this paper, we instead qualitatively assess whether the projections were sufficiently close to what transpired to be informative for policy.

There were a number of stages involved with scientific modelling for policy that operated throughout 2021 and the Roadmap for relaxation of controls. The modelling was coordinated through SPI-M-O, the Scientific Pandemic Influenza Group on Modelling, Operational subgroup of the Scientific Advisory Group for Emergencies (SAGE). Three main academic groups - from the University of Warwick, the London School of Hygiene and Tropical Medicine [2] and Imperial College London [9] - contributed model projections to most of the six Roadmaps. Preliminary results (one week before) and final results were presented at SPI-M-O meetings on Wednesdays to a large body of epidemiological modellers and other experts for comments and criticisms. The individual Roadmap documents from each institution, together with a comprehensive SPI-M-O summary document [20–25], were presented to SAGE the following day (Thursday). These projections together with epidemiological, behavioural and public health insights were then presented to government by the Chief Medical Officer and the Chief Scientific Advisor. This collection of advice, together with a range of other considerations, led to the final policy announcement on Monday - with the policy coming into force on the following Monday. The number of stages involved, passing through two expert committees and involving independent models, ensured some degree of robustness, but also added a considerable delay between the last available data on Monday or Tuesday evening and the announcement to the public, together with the release of the reports, on the following Monday.

The aim of each Roadmap document was to provide the government with an assessment of the likely implications of further future relaxations to the existing control measures. The first Roadmap document was very much focused on assessing the general time-scale of relaxation, which can be considered a balance between sufficient immunity from vaccination and greater freedom for increased social mixing. Roadmaps 2-4b, were concerned with the prescribed Steps 2-4, providing confidence to the government that none of the step-changes would overwhelm health-care resources. As a consequence, we frequently investigated a range of scenarios including more cautious assumptions about the action of vaccination or the response to each Step. Finally, Roadmap 5 was predominantly a look ahead to the 2021/2022 winter season, to assess whether a combination of seasonality and recently quantified waning immunity could generate public health issues. For each of the associated political decisions, such epidemiological projections are only one component and must be balanced against a range of social and economic factors.

While details of the model assumptions in each of the Roadmap documents differ, as both the situation and our understanding of the epidemiology changed, there are two main themes that are broadly consistent as time progresses. Firstly, as more data are collected there has been a general improvement in the accuracy of the vaccine related parameters that underpin each version of the model, often with a trend for the later, more refined estimates to suggest higher levels of vaccine protection. Secondly, during this process a better understanding of human behaviour in response to changing protective measures has been developed [26, 27]; in the early Roadmaps it was naively assumed that behaviour would track legal restrictions whereas population behaviour has proved to be far more complex. We introduce the models and results chronologically, comparing one of the main projection figures in each of the Roadmap documents [14–19] to assess whether the projections where sufficiently robust and accurate to help inform policy. As such, our aim is to bring together results generated in real-time during the pandemic and formulate lessons learned from this work.

## Results

The Roadmap results reflect key parameter assumptions and model structures which were based on the available data and information at the time. The Methods section provides details of the model formulation and assumptions for each Roadmap and how a range of parameters are inferred from the available epidemiological data.

We begin by considering the change in estimated level of precautionary behaviour over time (Fig. 1a). The precautionary behaviour parameter captures all social and behavioural changes that lead to a change in the underlying transmission rate (an increase in the level of precautionary behaviour leads to a decrease in transmission) - this includes enforced measures such as lockdowns and tiers, as well as voluntary measures such as avoiding crowded areas, mask wearing and lateral flow testing. Three key points can be observed from the inferred temporal pattern: (i) precautionary behaviour increases in response to imposed restrictions (such as the three lockdowns); (ii) there is a gradual decline in precautionary behaviour at other times, as individuals seek a return to normality during periods of relative stability; (iii) on top of this pattern there are numerous anomalies when the population responds to external events. For example, precautionary behaviour initially increased during June 2021 in response to concerns about the Delta variant, decreased dramatically in late June/July 2021 (during the 2020 UEFA European Football Championship), and then increased again due to increased isolation caused by the ‘pingdemic’.

**Fig. 1:**
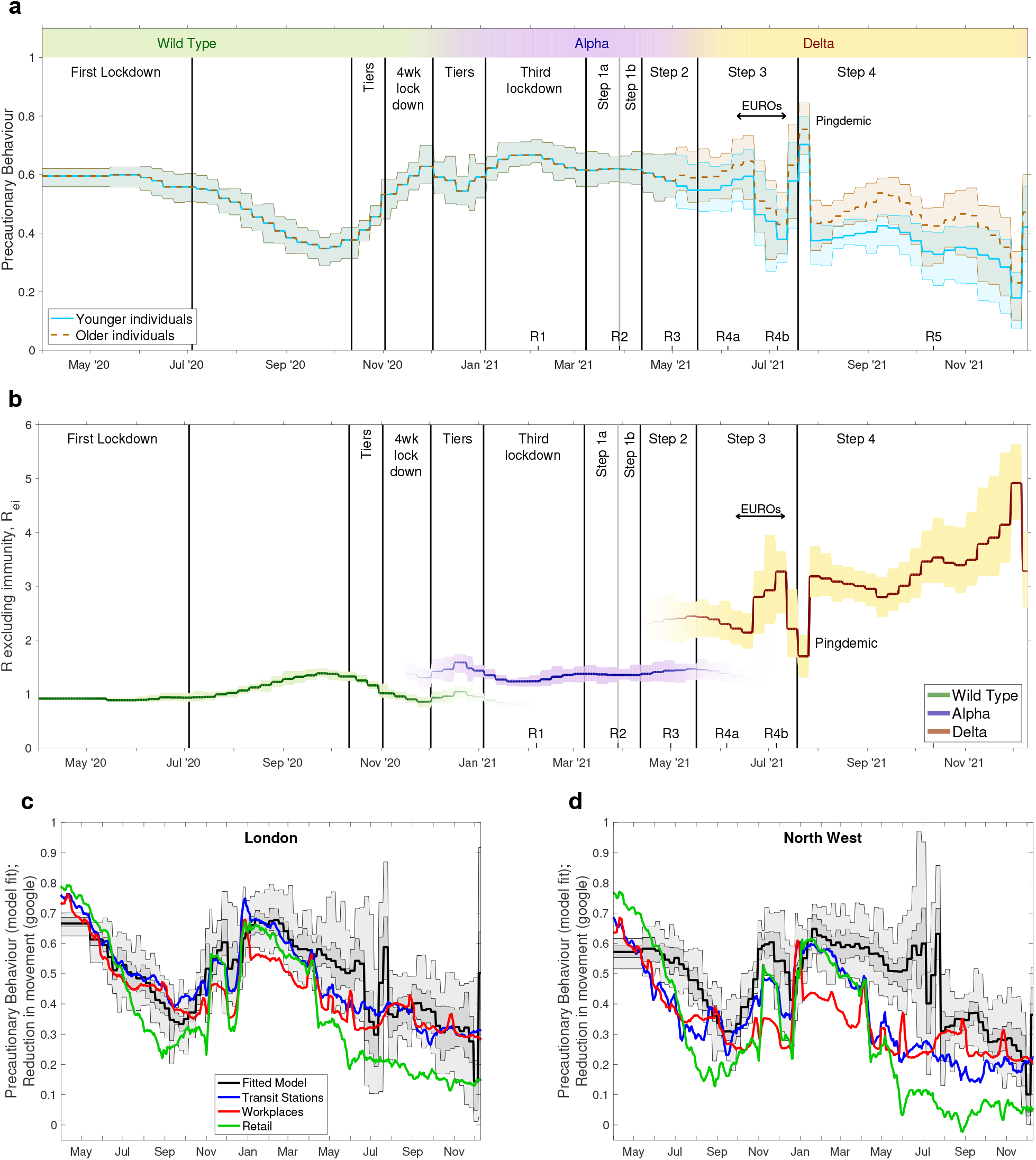
Changes in the inferred dynamics over time. **a** The inferred precautionary behaviour in England from April 2020 to December 2021; younger individuals are under 40, older individuals are over 65, while those in between are a linear extrapolation. **b** The resultant changes to *R* excluding immunity for the three main variants. **c, d** A comparison between fitted precautionary behaviour and Google movement records [49] in two NHS regions. Vertical lines indicate the time of key changes to the control measures. Labels are key moments from the pandemic in England: Lockdowns are nationwide restrictions on movement and social mixing; Tiers are localised restrictions of varying degrees; Steps are as described throughout this document; EUROS are the UEFA European Football Championships that led to substantially increased levels of social mixing; while the Pingdemic is a period when there was a vast increase in mobile-phone App alerts due to high levels of cases and more frequent social mixing. The dates of the six Roadmaps (R1-R5) are marked on the lower axes. In all panels the bold lines are mean values; in the top two panels (**a, b**), the shaded area represents the 95% credible interval, whereas in the bottom panels both the 50% (dark grey) and 95% (lighter grey) intervals are shown.

The inferred level of precautionary behaviour translates directly into the underlying transmission rate which, together with the impact of new variants, generates a reproductive ratio. We plot *R* excluding immunity (*R*_*ei*_), which captures what the reproductive ratio would be without the depletion of susceptibles due to prior infection or the actions of vaccination (Fig. 1b). As such *R*_*ei*_ provides an intuitive measure of the combined effects of behaviour and variants, and is directly inferred from precautionary behaviour and variant-specific parameters. Each variant has led to a noticeable rise in *R*_*ei*_, with Delta (and more recently Omicron) generating substantial increases over the initial wild type variant.

While the level of precautionary behaviour is inferred by matching to epidemiological data, it is interesting to compare the estimates to other measures of behaviour. In Figure 1c,d we compare our regional estimate of precautionary behaviour in London and the North West (two regions of England that have often suffered high infection burdens) with estimates of reduction in movement from Google mobility reports [49]. While we do not necessarily expect a one-to-one agreement, as the two statistics are measuring different quantities, there is striking similarity between our level of precautionary behaviour and Google’s measure of workplaces and transit stations in London. There is less correspondence in the North West, which could be attributable to a smaller proportion of the population using public transport (hence less accuracy for transit station measures) or greater control from measures unrelated to mass movement, such as greater testing and isolation or greater mask use.

Two key assumptions for each Roadmap document were the rate at which doses of vaccine could be delivered and the level of vaccine uptake within each age-group (Fig. 2). The expected level of uptake, together with assumptions about vaccine efficacy, defines the eventual impact of vaccination on the epidemic dynamics, while the deployment rate helps to define the time-scales over which this impact is achieved. Both the delivery rate and uptake quantities were fixed parameters in the default Roadmap modelling, based on information from the Cabinet Office, although we often considered sensitivity to these assumptions. Early Roadmap assumptions (Fig. 2, Roadmap 1 in red and Roadmap 2 in dark blue) were optimistic in terms of delivery speed, optimistic about uptake in the youngest age-groups but pessimistic about uptake in the older generations. Later Roadmaps were all slightly optimistic that vaccine delivery would be sustained at high levels for relatively long times, but included more accurate uptake assumptions. Roadmaps 1 to 4b did not include the vaccination of those under 18 nor the booster programme, hence these are not shown in Fig. 2; only Roadmap 5 includes these features as well as booster (third dose) vaccination.

**Fig. 2:**
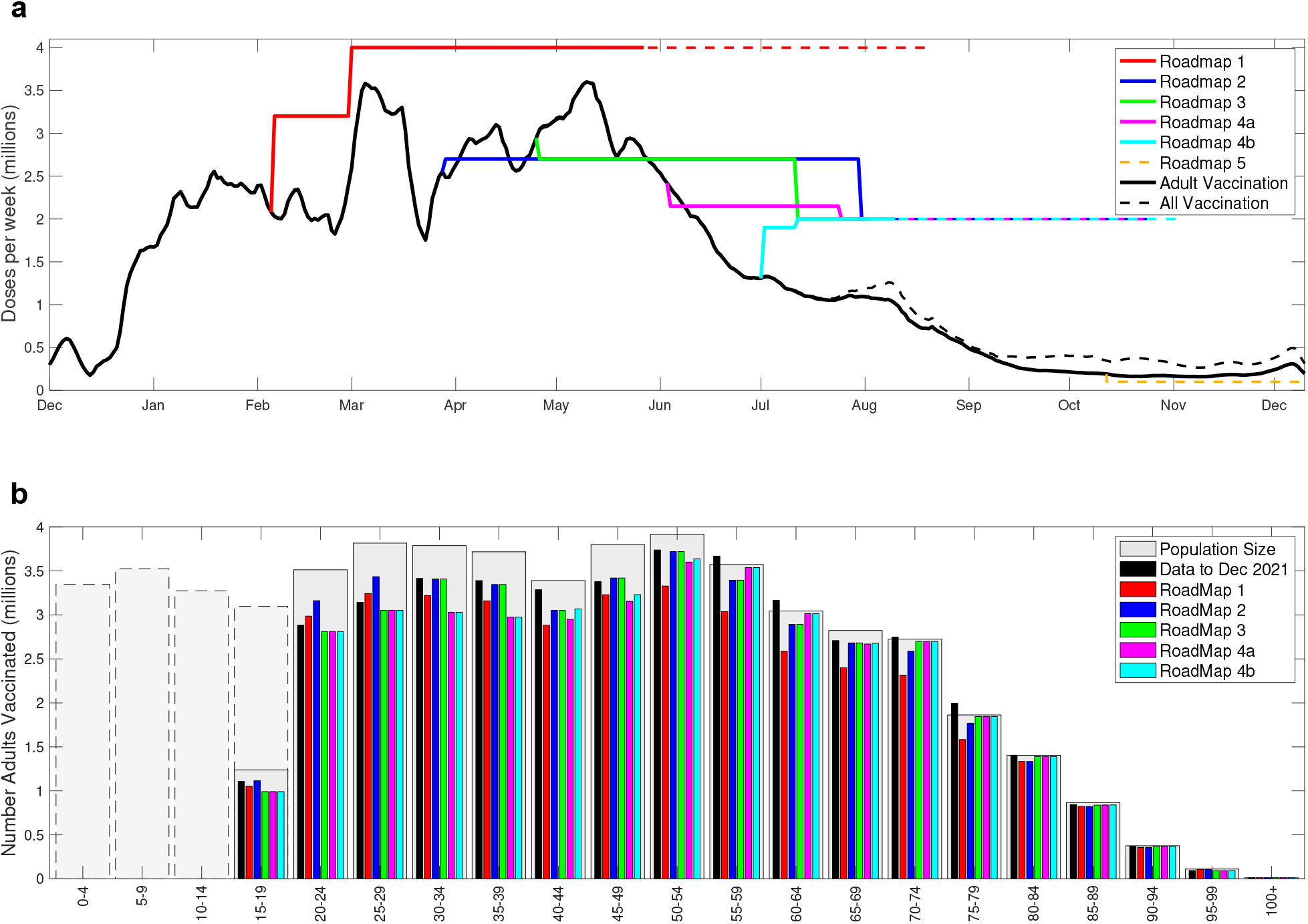
Assumptions about the vaccine uptake for the six roadmap models. **a** Realised delivery of vaccine in England, from December 2020 to December 2021 (black lines represent a 7-day moving average), compared with assumptions for the six Roadmap documents. The solid lines correspond to the time to give two doses to all adults (18+) at the assumed uptake; the black solid line is the reported rate of adult vaccination, while the black dashed line is the combined rate of vaccinating children and adults. **b** The resultant number of individuals eventually vaccinated in each 5-year age group. The pale grey bars represent the population size as estimated by ONS [50], with dashed borders representing individuals under 18 year of age. In the Roadmap 5 document, we based uptake on the observations taken in early October 2021.

### Roadmap 1, 6th February 2021

The earliest Roadmap was hampered by the limited amount of information in early February 2021 on the Alpha variant (especially in terms of vaccine protection) and a largely unknowable level of vaccine uptake. The assumed vaccine efficacy (based on trial data and early estimates in England) was slightly pessimistic, as was the assumed uptake level in older adults, but the rate of vaccine delivery was highly optimistic (Fig. 2a).

Figure 3 is taken from the February 2021 Roadmap document [14], plotting the projections for Scenarios Two, Three and Four, overlaid with the epidemiological data for England (up to December 2021). Considering the assumed changes in behaviour (Fig. 3a) it is clear that scenarios Three and Four, together with low transmission (generated through higher precautionary behaviour) in Step 5, are in closer agreement with the realised pattern of behaviour than the other assumptions considered; these correspond to the orange and yellow projections in the left-hand column of results (Fig. 3b-d).

**Fig. 3:**
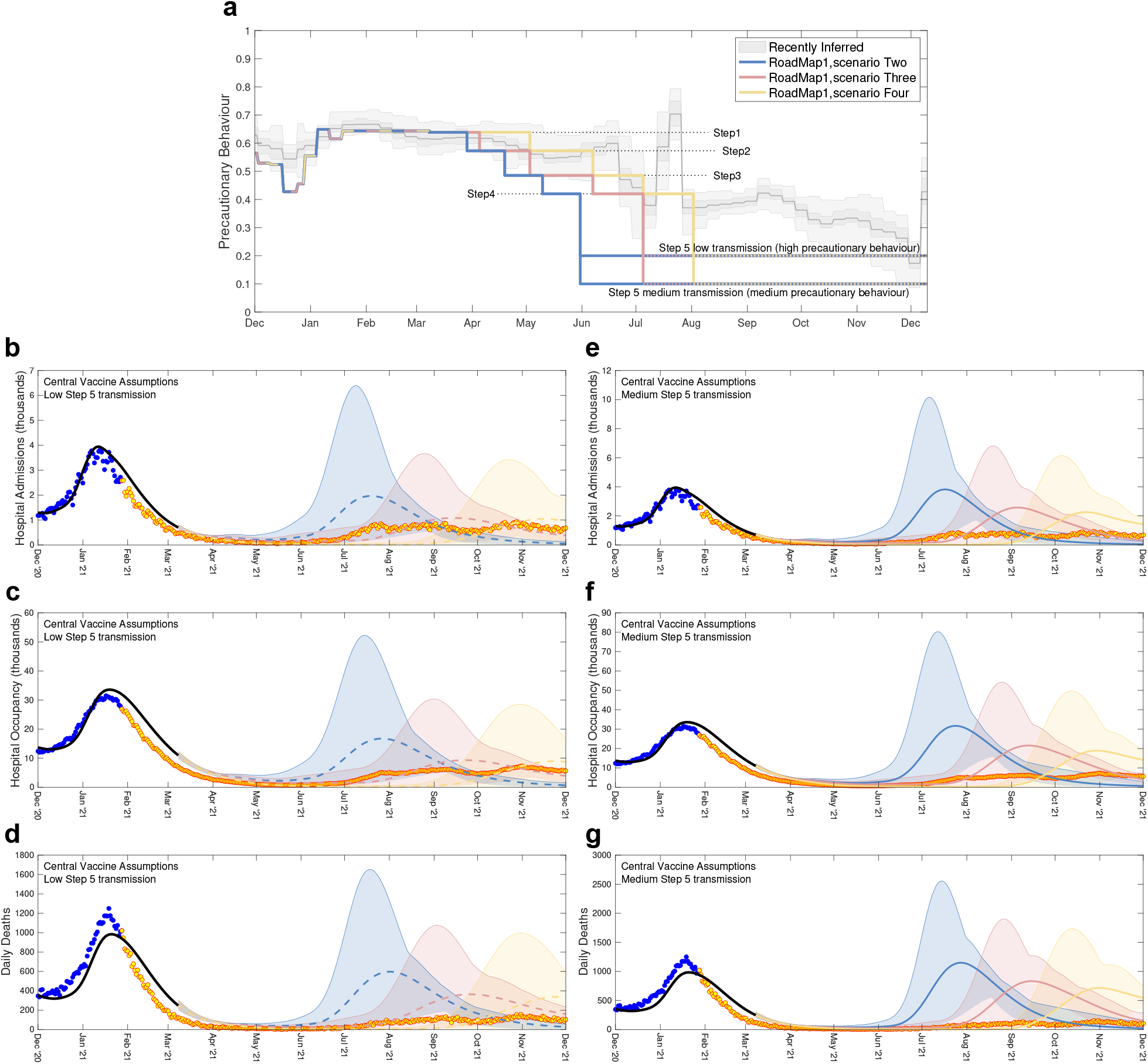
Severe disease projections from Roadmap 1 [14] together with precautionary behaviour assumptions. Low (**b-d**) and medium (**e-g**) transmission assumptions together with central vaccine efficacy assumptions are shown. **a** The most recently inferred level of precautionary behaviour (grey, showing mean, 50% and 95% credible intervals) together with three different scenarios and two levels of transmission in Step 5. The lower figures show projected level of daily hospital admissions (**b, e**), hospital occupancy (**c, f**) and daily deaths (**d, g**) together with the associated data until December 2021; data points from before the Roadmap and used in parameter inference are in blue, those after the Roadmap are in orange. Lines are the median values, while shaded regions correspond to the 95% prediction interval; blue for Scenario Two, orange for Scenario Three and yellow for Scenario Four.

The data for the latter half of 2021 lie well within the 95% prediction intervals (shaded regions) of the central vaccine efficacy assumptions, which are wide due to the considerable uncertainty of longterm projections. For hospital admissions and occupancy the projected median trajectories provide a reasonable approximation to the scale of outbreak. The model results do however over-estimate the number of deaths; the lower realised number of deaths may be attributable to better treatments administered in 2021 lowering the case-fatality ratio [51]. Four factors influence the comparison between model and data: (i) the model does not include the Delta variant; (ii) the assumed precautionary mixing in Step 5 was lower than inferred; (iii) the initial estimates of vaccine protection were too low (as shown in Table 1); and (iv) the forecast deployment speed of vaccination was higher than observed (Fig. 2a). The actions of (ii) and (iii) are to push the projection to be an overestimate of the true disease severity, while (i) and (iv) would generate underestimates. Our failure to predict the long-term plateau of severe disease through September and October 2022 is largely attributable to the emergence of the Delta variant, with higher transmission, the slow relaxation of population behaviour and the gradual waning of vaccine-derived protection; this is discussed in more detail in Roadmap 4b.

**Table 1:** Assumed percentage protection from vaccination against Alpha and Delta variants from the six Roadmap documents. For Roadmap 1 [14] Cautious/Central assumptions are shown; for Roadmap 4a [17] Cautious/Central/Optimistic assumptions are shown; for Roadmap 5 [19] we also include the level of waning immunity (where protection wanes to one of three values corresponding to a half-life of 180/270/460 days) and the action of booster vaccines (two assumptions for short-term and long-lasting immunity respectively) and. All values are based on parameters that were available at the time [33–40, 43–48], for the Pfizer-BioNTech BNT162b2 vaccine (Pf) and for the Oxford-AstraZeneca ChAdOx1 vaccine (AZ); the Moderna vaccine (Spikevax) is assumed to have the same efficacy as the Pfizer-BioNTech vaccine.

| Variant | Protection against | Vaccine | Dose | Roadmap |  |  |  |  |  |  |  |
| --- | --- | --- | --- | --- | --- | --- | --- | --- | --- | --- | --- |
|  |  |  |  | 1 | 2 | 3 | 4a | 4b | 5 | waning | booster |
| Alpha | Infection | AZ | 1 | 24/48 | 60 | 60 | 60/63/70 | 63 | 63 | - | - |
|  |  |  | 2 | 30/60 | 65 | 65 | 70/78/90 | 78 | 78 | 0/30/50 | - |
|  |  | Pf | 1 | 24/48 | 60 | 60 | 55/63/70 | 63 | 63 | - | - |
|  |  |  | 2 | 30/60 | 85 | 85 | 70/80/90 | 80 | 80 | 0/30/50 | - |
|  | Symptoms | AZ | 1 | 56/70 | 60 | 60 | 55/63/70 | 63 | 63 | - | - |
|  |  |  | 2 | 70/88 | 70 | 80 | 70/86/90 | 80 | 80 | 10/35/55 | - |
|  |  | Pf | 1 | 56/70 | 60 | 60 | 55/63/70 | 63 | 63 | - | - |
|  |  |  | 2 | 70/88 | 90 | 90 | 85/88/90 | 88 | 88 | 10/35/55 | - |
|  | Hospital Admission | AZ | 1 | 56/70 | 80 | 80 | 75/80/85 | 80 | 80 | - | - |
|  |  |  | 2 | 70/88 | 90 | 90 | 90/93/95 | 90 | 90 | 70/79/85 | - |
|  |  | Pf | 1 | 56/70 | 80 | 80 | 75/80/85 | 80 | 80 | - | - |
|  |  |  | 2 | 70/88 | 90 | 90 | 90/93/95 | 93 | 93 | 70/79/85 | - |
|  | Mortality | AZ | 1 | 56/70 | 80 | 80 | 75/78/80 | 80 | 80 | - | - |
|  |  |  | 2 | 70/88 | 90 | 90 | 95/97/99 | 95 | 95 | 70/79/85 | - |
|  |  | Pf | 1 | 56/70 | 80 | 80 | 75/78/80 | 80 | 80 | - | - |
|  |  |  | 2 | 70/88 | 90 | 90 | 95/97/99 | 97 | 97 | 70/79/85 | - |
|  | Onward Transmission | AZ | 1 | 0 | 0 | 40 | 40/45/50 | 45 | 45 | - | - |
|  |  |  | 2 | 0 | 0 | 50 | 40/45/50 | 45 | 45 | 20 | - |
|  |  | Pf | 1 | 0 | 0 | 50 | 40/45/50 | 45 | 45 | - | - |
|  |  |  | 2 | 0 | 0 | 50 | 40/45/50 | 45 | 45 | 20 | - |
| Delta | Infection | AZ | 1 | - | - | - | 33/34/45 | 34 | 45 | - | - |
|  |  |  | 2 | - | - | - | 58/71/84 | 64 | 70 | 0/30/50 | 85/92 |
|  |  | Pf | 1 | - | - | - | 30/34/45 | 56 | 55 | - | - |
|  |  |  | 2 | - | - | - | 58/73/84 | 80 | 85 | 0/30/50 | 85/92 |
|  | Symptoms | AZ | 1 | - | - | - | 27/34/45 | 34 | 45 |  |  |
|  |  |  | 2 | - | - | - | 64/82/84 | 70 | 70 | 10/35/55 | 90/95 |
|  |  | Pf | 1 | - | - | - | 24/34/45 | 56 | 55 | - | - |
|  |  |  | 2 | - | - | - | 64/83/84 | 88 | 90 | 10/35/55 | 90/95 |
|  | Hospital Admission | AZ | 1 | - | - | - | 60/64/85 | 81 | 80 | - | - |
|  |  |  | 2 | - | - | - | 86/90/95 | 94 | 95 | 70/79/85 | 95/97 |
|  |  | Pf | 1 | - | - | - | 58/64/85 | 90 | 80 | - | - |
|  |  |  | 2 | - | - | - | 86/91/95 | 98 | 95 | 70/79/85 | 95/97 |
|  | Mortality | AZ | 1 | - | - | - | 60/60/80 | 81 | 80 | - | - |
|  |  |  | 2 | - | - | - | 93/96/99 | 95 | 98 | 70/79/85 | 98/99 |
|  |  | Pf | 1 | - | - | - | 58/60/80 | 90 | 80 | - | - |
|  |  |  | 2 | - | - | - | 93/96/99 | 98 | 98 | 70/79/85 | 98/99 |
|  | Onward Transmission | AZ | 1 | - | - | - | 40/45/50 | 45 | 30 | - | - |
|  |  |  | 2 | - | - | - | 40/45/50 | 45 | 30 | 20 | 30 |
|  |  | Pf | 1 | - | - | - | 40/45/50 | 45 | 30 | - | - |
|  |  |  | 2 | - | - | - | 40/45/50 | 45 | 30 | 20 | 30 |

### Roadmap 2, 29th March 2021

Roadmap 2 considered two distinct scenarios: the impact of just allowing Step 2 (opening of nonessential retail and outdoor hospitality venues); and the longer term dynamics of Steps 2-4. For the longer term projections, the Roadmap document [15] considered a number of different sensitivities in order to explore a range of parameter uncertainties including: the strength of social mixing (and hence transmission) in Step 4; the strength of social mixing in Steps 2 and 3; seasonality, such that transmission is lower in the summer months; as well as vaccine roll-out speed, efficacy and uptake. Of these uncertainties we focus here on the inclusion of seasonal effects (Fig. 4), as we subsequently included moderate levels of seasonality (10%) in all future Roadmaps. Here, seasonality is a measure of the peak-to-trough drop in transmission relative to the peak; we captured this process by a sinewave, whose trough was centred on mid-August based on changes in relative humidity [52], which also provide the justification for the 10% seasonality used in subsequent Roadmaps for England [52].

**Fig. 4:**
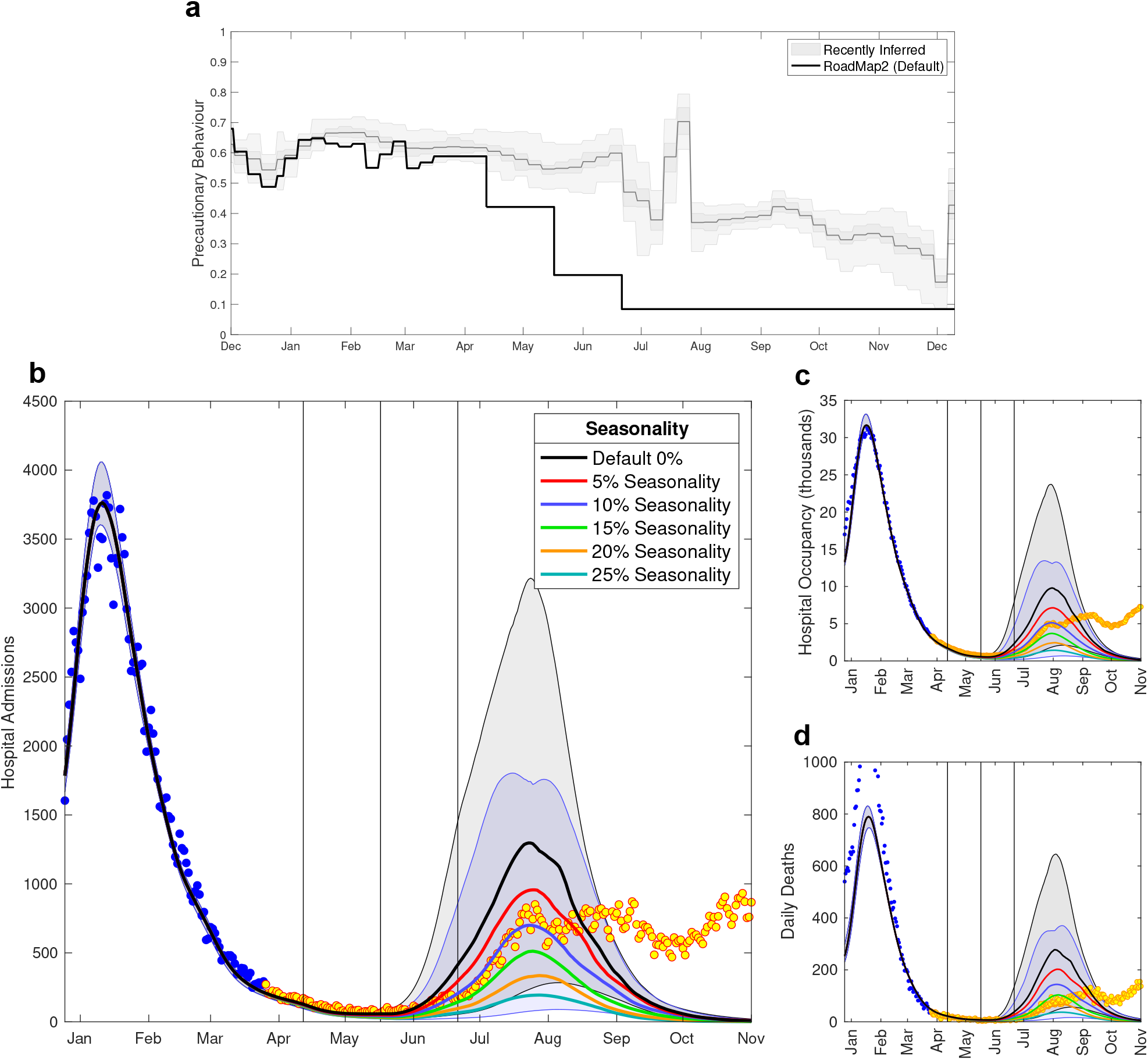
Severe disease projections from Roadmap 2 Figure 2.8 [15], which considered the impact of seasonality. **a** The most recently inferred level of precautionary behaviour (grey, showing mean, 50% and 95% credible intervals) together with the assumed default levels from Roadmap 2 [15]. Lower panels show the model outputs for daily hospital admissions (**b**), hospital occupancy (**c**) and daily deaths (**d**) together with the associated data until December 2021; data points from before the Roadmap and used in parameter inference are in blue, those after the Roadmap are in orange. Lines are the median values, while shaded regions correspond to the 95% prediction interval, and are only shown for the default scenario (no seasonality, black) and 10% seasonality scenario (blue) that is used in later models. Vertical lines are the timing of Steps 2-4.

Roadmap 2 overestimated the degree to which the population would return to pre-COVID mixing in each Step of restrictions being eased (Fig. 4a). Additionally, the Roadmap 2 document only considered the dynamics of the Alpha variant, whereas in May 2021 the Delta variant invaded increasing *R* by an estimated 67% (CI 44-95%) (Fig. 1b). The action of these two factors works in opposite directions: our assumed lower level of precautionary behaviour will act to increase our projections, while maintaining the Alpha variant will suppress the projections.

We highlight outcomes under different seasonality assumptions. In particular, we investigated the impact of assuming no seasonality (Fig. 4, black line) and 10% seasonality (as assumed in future Roadmaps, represented in Fig. 4 by the blue line), both of which are shown with 95% prediction intervals (shaded regions). The data on hospital admissions (Fig. 4b), hospital occupancy and deaths (Fig. 4c,d) all fall within the 95% prediction interval of 0% and 10% seasonality until late August 2021; while the assumption of 10% seasonality leads to an excellent agreement between the median prediction and the hospital data prior to the peak in late July 2021. The median deaths are again an overestimate, likely due to improvements in medical treatments that were applied as we progressed through 2021. In addition, we included in later Roadmaps a correlation between the mortality risk and hospital occupancy, meaning smaller waves in the summer of 2021 would be associated with relatively lower deaths than the major wave of January 2021.

### Roadmap 3, 4th May 2021

Our assumptions in Roadmap 3 [16] about social mixing associated with Steps 3 and 4 significantly underestimated the level of precautionary behaviour (Fig. 5a), leading to modelled outputs with greater transmission than was realised. Even our low transmission assumption (blue line) was an underestimate of precautionary behaviour from late May until December 2021. Despite this, our projections of hospital admissions and hospital occupancy were underestimates of the true dynamics (Fig. 5b,c) – although the majority of the data points until September 2021 fall within the 95% prediction intervals. This is most clearly seen during May and early June 2021 when hospital admissions began to slowly climb, but the median projection was for relatively constant levels of hospital admissions until the impact of Step 4 was realised.

**Fig. 5:**
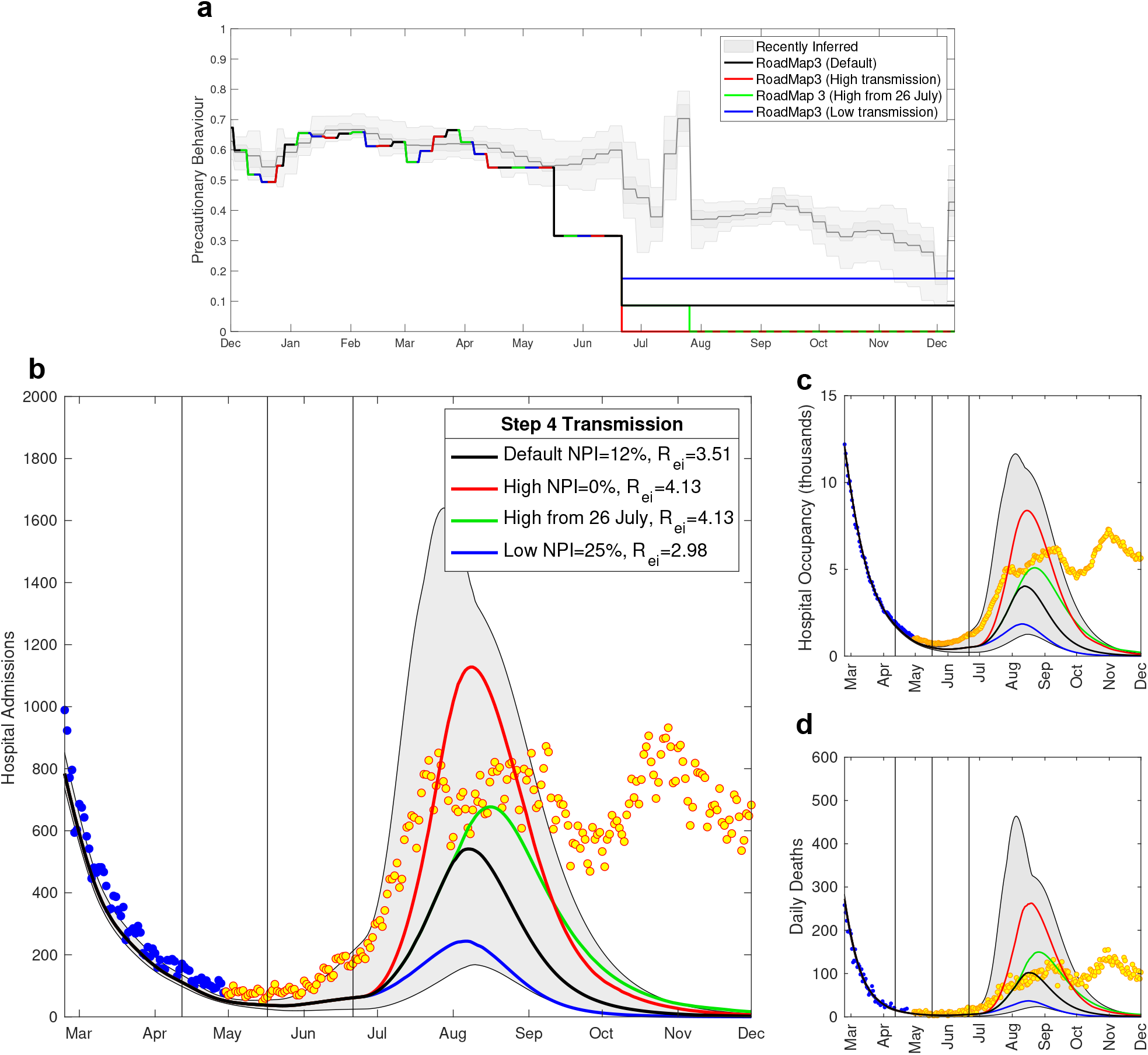
Severe disease projections from Roadmap 3 Figure 10 [16], which considered the impact of social mixing and hence transmission in Step 4. **a** The most recently inferred level of precautionary behaviour (grey, showing mean, 50% and 95% credible intervals) together with the assumed default levels from Roadmap 3 [16]. Lower panels show projected level of daily hospital admissions (**b**), hospital occupancy (**c**) and daily deaths (**d**) together with the associated data until December 2021; data points from before the Roadmap and used in parameter inference are in blue, those after the Roadmap are in orange. Lines are the median values, while the shaded region correspond to the 95% prediction interval for the default model (black); vertical lines correspond to the expected date of Steps 2, 3 and 4 on 12th April 2021, 17th May 2021 and 21st June 2021.

There are two main reason for this underestimate. Firstly, given we expected *R* to be close to one, even relatively small errors can translate into substantial divergence in projections - changing trajectories from flat to exponentially increasing. However, the key driver is undoubtedly the invasion and increase of the Delta variant in late May 2021. Subsequent analysis suggests that Delta is 67% (CI 44-95%) more transmissible than Alpha [19], a factor that is more than sufficient to overcome the errors in our assumptions about precautionary behaviour and drive pronounced exponential growth.

### Roadmap 4a, 8th June 2021

The assumptions in Roadmap 4a [17] for Step 3 and 4 clearly underestimated the level of precautionary behaviour (Fig. 6a). The heightened concerns in late May and early June 2021 over the invasion of the Delta variant led to an increase in precautionary behaviour at a time when we were anticipating a decline due to the relaxation of Step 3. Again, the level of social mixing assumed in Step 4 is far higher (analogously the precautionary behaviour was far lower) than observed at any point in 2021. In particular, the ‘pingdemic’ of late July 2021 which led to a massive rapid decline in infection was not included in the modelled scenarios and could not have been forecast. With the mounting concern over the Delta variant, one of the pivotal considerations of Roadmap 4a is whether Step 4 (originally scheduled for 21st June 2021 at the earliest) should be delayed. Given the substantial uncertainty in estimating vaccine efficacy against Delta, this Roadmap considered three assumptions about vaccine protection: default, optimistic and cautious (Fig. 6, b-d). Later studies (see Table 1) suggest that the optimistic assumptions are closest to the level of protection realised. The default assumptions therefore, unsurprisingly, overestimate the true level of hospital admissions; the model projected a far more rapid rise in the Delta variant due to less suppression by vaccine immunity and no public reaction to the new variant. When using the optimistic vaccine assumptions (lower left panel), there is closer correspondence between the model and how the outbreak unfolded: the data comfortably lie within the 95% prediction interval until September 2021 for the realised scenario of Step 4 on 19th July 2021 (Fig. 6c, orange line and shaded area).

**Fig. 6:**
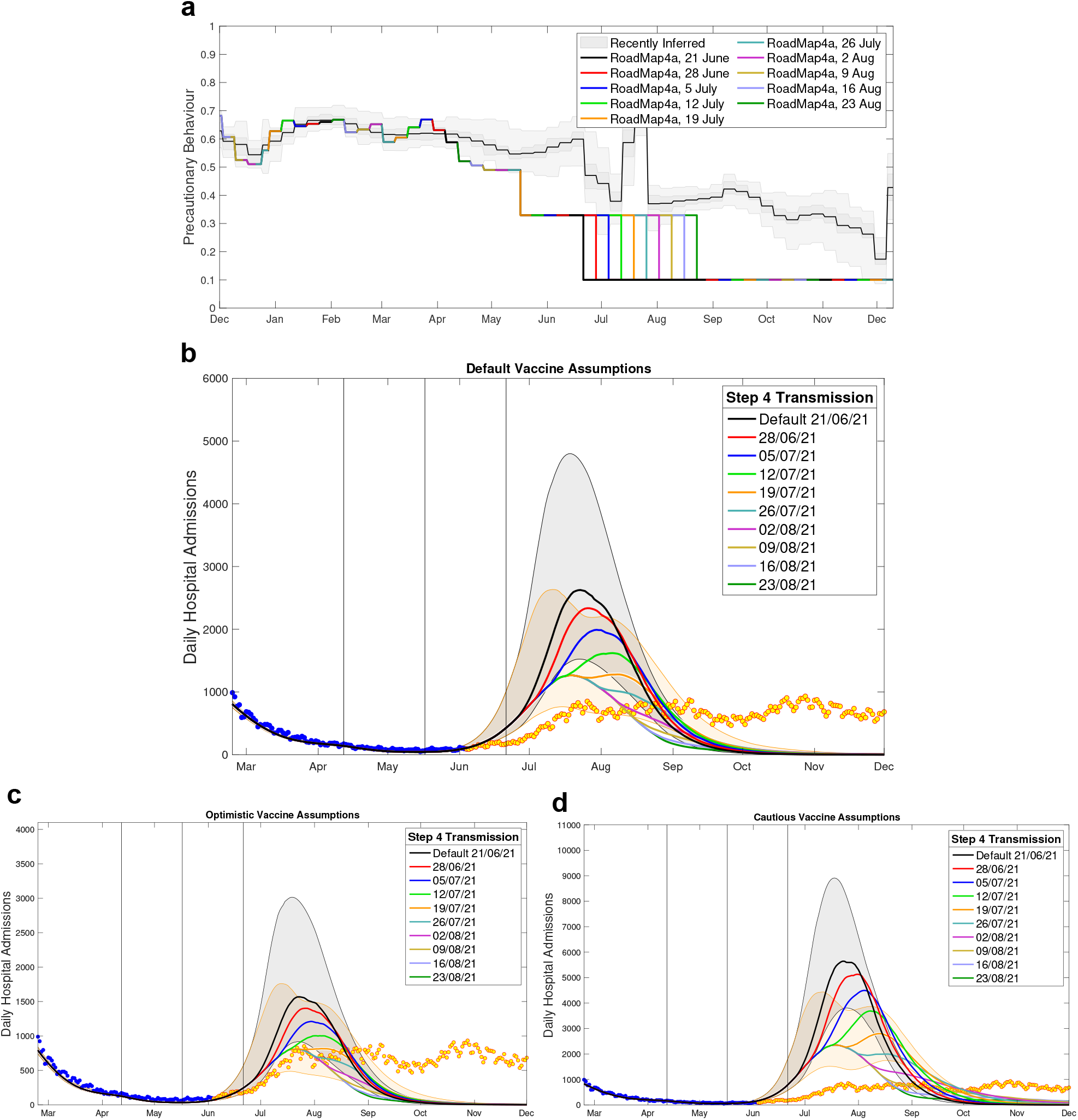
Hospital admission projections from Roadmap 4a Fig. 10 [17], which considered the impact of delaying the transition to Step 4. **a** The most recently inferred level of precautionary behaviour (grey, showing mean, 50% and 95% credible intervals) together with the assumed changes in Roadmap 4a at a range of assumed delays for Step 4. **b-d** projected level of daily hospital admissions for the default (**b**), optimistic (**c**) and cautious (**d**) assumptions about vaccine protection, together with the associated data until December 2021; data points from before the Roadmap and used in parameter inference are in blue, those after the Roadmap are in orange. Lines are the median values, while shaded regions correspond to the 95% prediction intervals for the default model (black) and the delay until 19th July 2021 (orange).

It could be questioned why Roadmap 4a predicted a single, although broad-peaked, epidemic, while in reality the Delta variant maintained a plateau of hospital admissions for a duration of around four months. Again, this is driven by the underlying estimates of precautionary behaviour. In particular, we had difficulty estimating the change due to Step 3, which was exacerbated by the emergence of the Delta variant, the ensuing public response to this threat and the short time between Step 3 and producing Roadmap 4a. Any substantial drop in precautionary behaviour is likely to drive a rise in cases (and hence a rise in hospital admissions) which will eventually decay, leading to a single-humped wave. In contrast, what was observed was a complex pattern of falls and rises, followed by a more gradual decline in precautionary behaviour as people reacted to changing risk (as exemplified by [53]), all of which led to the long high plateau of hospital admissions during the Delta wave. In addition, the waning of vaccine efficacy, especially in the elderly, who were amongst the first to be vaccinated, also acted to promote the high plateau of hospital admissions.

### Roadmap 4b, 6th July

The assumptions in Roadmap 4b [18] for the behaviour in response to moving to Step 4 cover a wide range of scenarios (Fig. 7, top panel); however, none captured the sudden increase in mixing during the 2020 UEFA European Football Championships, nor the sudden subsequent decline from the ‘pingdemic’ (a sudden surge in contact tracing App alerts due to a combined increase in mixing and cases). With the advantage of hindsight, the fact that these events would precipitate changes may have been anticipated but their scale was unforeseeable. Of the scenarios considered, the slow decline (Fig. 7a, green line) was the closest to the realised pattern of social mixing but, since there were pronounced changes in behaviour during June and July 2021 that were absent from the modelled scenarios, we would not expect the associated modelled projections to closely match the data.

**Fig. 7:**
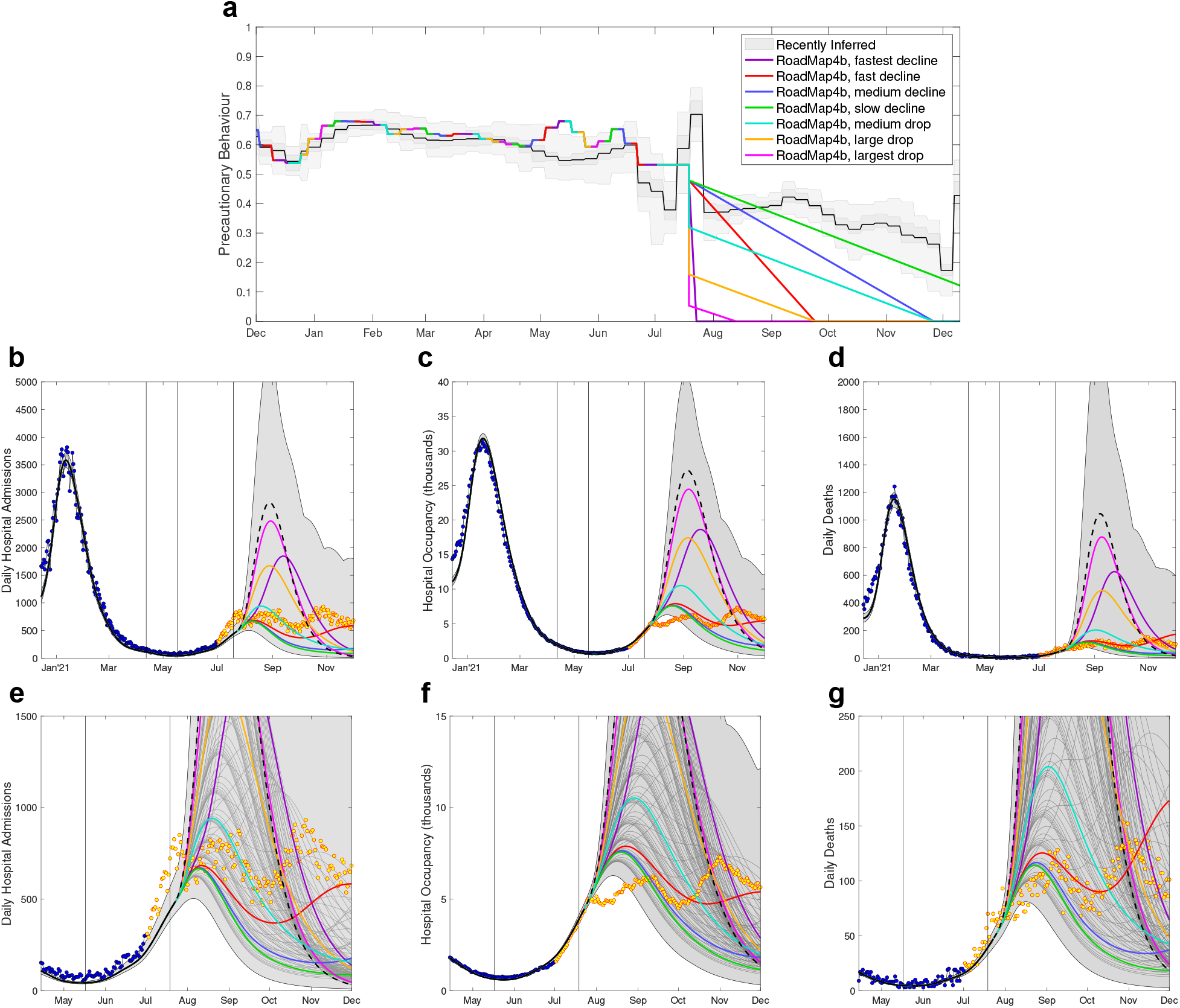
Severe disease projections from Roadmap 4b Figure 5 [18], which shows the projections associated with seven assumptions about Step 4 precautionary behaviour. **a** The most recently inferred level of precautionary behaviour (grey, showing mean, 50% and 95% credible intervals) together with the seven different assumptions from Roadmap 4b. The lower panels show daily hospital admissions (**b**,**e**), hospital occupancy (**c**,**f**) and daily deaths (**d**,**g**) for the seven main assumptions (coloured lines giving the median values) and for the naive assumption of an abrupt return to pre-COVID mixing (black dashed line) together with the combined envelope of all 95% prediction intervals (shaded grey). The lower panels (**e-g**) show a magnified section of upper panels (**b-d**), which include median projections for other intermediate declines in precautionary behaviour (thin dark-grey lines). Data points from before the Roadmap and used in parameter inference are in blue, with subsequent data points displayed in yellow. Vertical lines show the dates of Steps 2 and 3, and the revised Step 4.

The data predominantly lie within the envelop of model projections. Many of the individual median trajectories (for example the green, blue, red and cyan trajectories Fig. 7, b-g) capture the approximate level of severe disease – although in relative terms hospital occupancy is lower than hospital admissions due to a slight reduction in the average length of stay in hospitals over this period. The data between July 2021 and December 2021 is characterised by a high plateau of infection with gradual oscillations that we conjecture is caused by a finely balanced combination of susceptible depletion, waning immunity and changes in social mixing. As such, many of the median trajectories (e.g. red curve) show this type of pattern, which comes from over-compensatory cycles (of the kind that generate traditional damped oscillations around the endemic equilibrium) with the linear decline in precautionary behaviour maintaining a high level of infection, whereas a constant level of precautionary behaviour would lead to a single-humped wave. However, we should not expect a close match between the data and any one trajectory, as the perturbations that occurred in June and July 2021 will have long-lasting consequences by changing the level of infection and the number of susceptibles.

### Roadmap 5, 12th October

This final Roadmap of 2021, considering possible SARS-CoV-2 transmission dynamics and the associated COVID-19 disease burden for Autumn and Winter 2021, was by far the most complex in terms of the dynamics being modelled. The Roadmap document also sought to capture the widest range of uncertainties in combination: three assumptions about waning immunity (red, green and blue in Fig. 8), two assumptions about the duration of boosters (centre and right-hand panels in Fig. 8), and multiple assumptions about the return to pre-pandemic mixing (of which we show three: cyan, magenta and purple in Fig. 8 rows b-d).

**Fig. 8:**
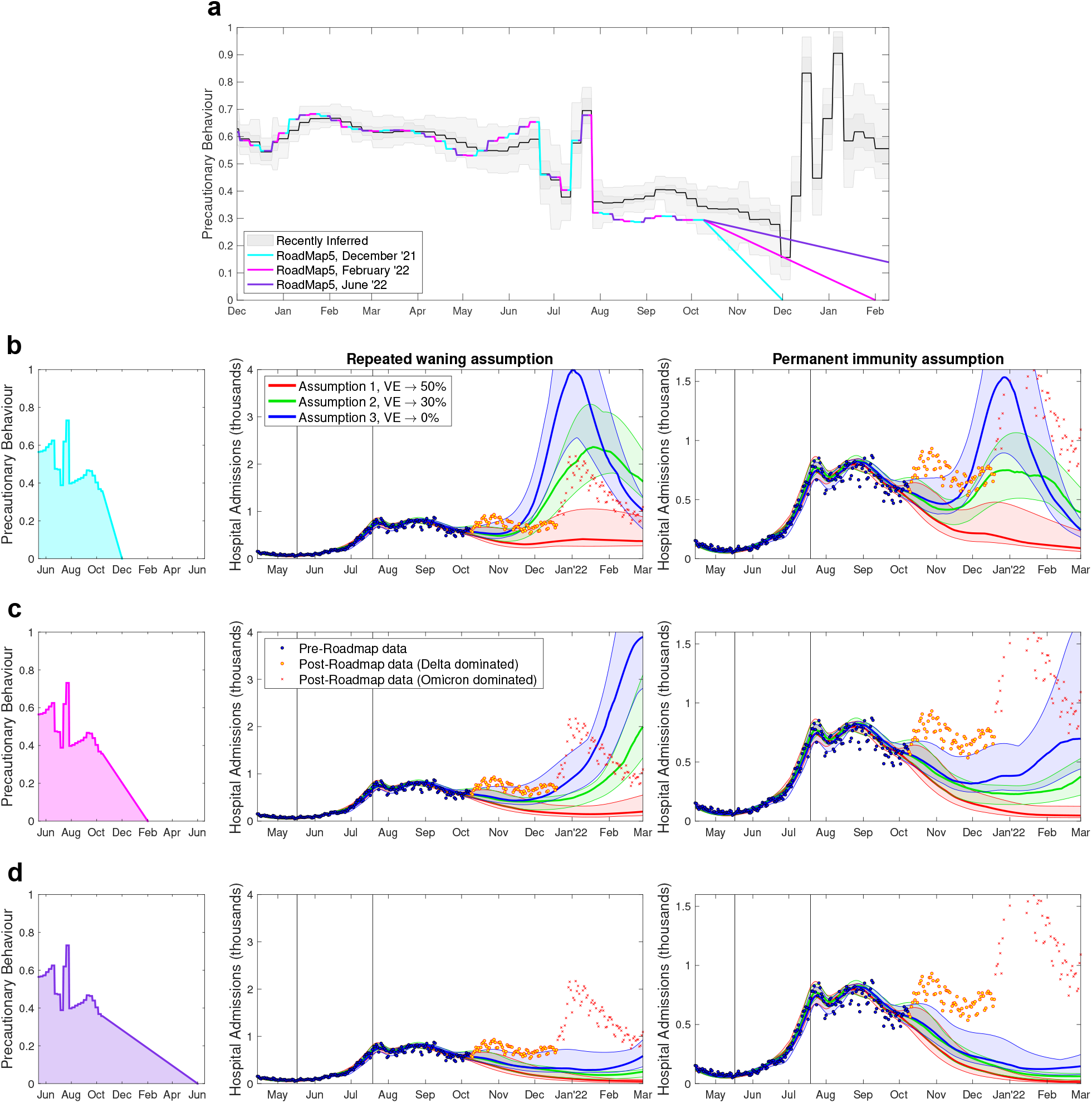
Hospital admission projections from Roadmap 5 Fig. 5 [19]. Scenarios cover three assumptions about precautionary behaviour (rows **b-d**), three assumptions about waning vaccine efficacy (red, green blue) and two assumptions about the long-term action of boosters (repeated waning in centre column and long term protection in the right column). **a** the most recently inferred level of precautionary behaviour (grey, showing mean, 50% and 95% credible intervals) together with the three different assumptions (cyan, pink and purple) about the return to pre-pandemic mixing used in Roadmap 5. Rows **b**-**d** show daily hospital admissions for the different assumptions (solid lines are the medians and shaded regions are the 95% projection intervals); points correspond to the data, with marker style denoting the times corresponding to dates before the projections were performed (blue circles), later times dominated by Delta (orange circles) and later times dominated by Omicron (red crosses). Vertical lines show the dates of Step 3 and the revised Step 4.

The four assumptions about the return to pre-pandemic mixing (Fig. 8a) clearly span the range of inferred behaviour, although our initial estimates of precautionary behaviour in August and September 2021 were lower than our most recent estimates for these values– with implications for the inferred parameters for the Delta variant (including the transmission rate and the risk of requiring hospital treatment).

The projections show the true number of hospital admissions in England both when the Delta variant dominated (orange circles, Fig. 8b-d) and at later times when the Omicron variant generated the majority of hospital admissions (red crosses, Fig. 8b-d). Although the model fits well to the data available at the time of simulation, it fails to capture the final oscillatory rise in Delta cases (during October 2021), and as expected cannot predict the Omicron wave. This discrepancy has three main attributes: the absence of a clear signal of another rise in the data at the time the inference and simulations were performed; a slight but systematic bias in the estimation of precautionary behaviour during much of the Delta wave; and a lack of detailed quantitative information on waning immunity and the long-term protection against hospital admissions. We address these three points in turn. First, without a clear signal of rising cases or rising hospital admissions, the model (unsurprisingly) continues the most recent trend of declining admissions throughout October 2021; in a relatively short period where frequent oscillations are observed it is difficult to identify such patterns in the noisy data. Second, the perturbations due to the UEFA European Football Championships in June 2021 and the pingdemic in July 2021, together with problems with testing in September and October 2021 [54] caused substantial difficulties is estimating the level of precautionary behaviour. Finally, the most recent data suggest a notable difference in waning between AstraZeneca and Pfizer vaccines in terms of protecting against symptomatic infections (closer to Assumption 3, blue curve in Fig. 8b-d), although both vaccines maintain relatively high levels of protection against hospital admissions (closer to Assumption 1, red curves in Fig. 8b-d).

## Discussion

Here we have discussed the development of the model outputs, generated using the Warwick SARS-CoV-2 transmission and COVID-19 disease model, that underpinned the six Roadmap documents produced to study the easement of restrictions in England during 2021. The chronological evolution across these documents is characterised by an increased understanding of vaccine efficacy, a greater appreciation for the uncertainty in population behaviour and a corresponding increase in model complexity. In general we believe the models acted as a useful policy tool; although the median projected wave of hospital admissions never precisely matched the realised data (sometimes over-estimating, sometimes under-estimating) there was in general an overlap between the data and the 95% prediction intervals over multiple models. In respect of the aim to provide scientific evidence before each step in the relaxation of controls [11], suggesting when the steps would lead to a minor increase in cases or when greater caution may be needed, in our view the six Roadmap documents fulfilled their purpose, although not without their critics [10, 55]. The work highlights two clear lessons for the future: firstly, the difficulties of making detailed long-term predictions due to uncertainties in vaccine efficacy (until large amounts of data are collected), the uncertainty in human behaviour, intrinsic uncertainties in the data, and the unknowable timing of any new variant of concern; and secondly, the difficulties of communicating projections to a policy and lay audience. We deal with these issues in turn.

The six Roadmap models have been driven by externally generated estimates of vaccine efficacy (Table 1). Any changes to these assumptions requires a refitting of the model to the entire period since vaccination began, and therefore we were constrained to use a small number of vaccine assumptions. Moreover, small changes to vaccine efficacy can generate large changes in model outcomes; changing the vaccine efficacy against death from 98% to 99%, for example, would halve the number of COVID-19 related mortalities in the vaccinated group. Similarly, small changes in the vaccine efficacy against infection or the reduction in onward transmission (both of which are inherently difficult to measure directly) will generate changes in growth rate; long-term projections are highly sensitive to any variation in the growth rate due to the amplifying effects of exponential processes. In general, early estimates of vaccine efficacy have been slightly pessimistic (comparing early and later values in Table 1), which may be due to early biases in the initial vaccine prioritisation as well as biases due to targeting those most likely to contract SARS-CoV-2 and require treatment.

A second consistent issue is estimating population behaviour in response to changes in legislation, case numbers and media messaging. Although we often conceptualise precautionary behaviour in terms of social mixing, it really refers to a reduction in risky mixing between susceptible and infected individuals. Therefore, mass testing followed by strict adherence to isolation rules is likely to prevent more transmission and hence lead to a higher estimate of precautionary behaviour than a simple change in the population-wide level of social mixing. Similarly, mask wearing will also reduce transmission and hence be reflected as an increase in precautionary behaviour.

The initial models [14–17], in the absence of any other data, assumed that changes in legislation would generate a step-change in population behaviour as people took advantage of new freedoms and opportunities. While the assumption of a step-change responding to relaxing measures was supported by the sudden increases in precautionary behaviour when measures were initially introduced (Fig. 1), we now observe that the response to the introduction of measures was much quicker than the response to the relaxation of measures. While with hindsight it is clear that behaviour was only responding slowly to each Step, in real time and with a variety of influencing factors this was difficult to detect; for example, in Roadmap 4a it had been impossible to robustly determine the impact of Step 3 (due to insufficient data and the complicating spread of the Delta variant), and the inferred change following Step 2 was relatively close our earlier assumptions.

Later models [18, 19] assumed a more gradual response to the relaxation steps; but population behaviour remains unpredictable over long timescales. Although we have generally seen a decline in the inferred precautionary behaviour (and hence an increase in social mixing) since the end of the January 2021 lockdown (Fig. 1) the change has been anything but smooth. While an increase in mixing during the 2020 UEFA European Football Championship, which occurred in late June to early July 2021, could have been foreseen, the magnitude and duration was unpredictable; similarly the ‘pingdemic’ of late July 2021 was an unexpected and emergent phenomenon that only subsided due to a change in the NHS App (switching from a notification window of 5 days to just 2 days for asymptomatically infected individuals, and hence minimising the number of individuals isolating). As such, the relaxation of legislation for each Step acts to provide a lower bound for precautionary behaviour by placing strict limits on social mixing, but multiple factors that contribute to individual perceived risk determine the true level. In December 2021, warnings and concerns over the Omicron variant generated marked changes in behaviour over the Christmas and New Year period (Fig. 8a) in the absence of legislation. The challenges posed in bringing together behavioural science and epidemiological theory have been recognised for some time in the infectious disease modelling community [56]. That being said, recent theoretical modelling analysis that have coupled transmission dynamics to behavioural mechanisms have illustrated emergent phenomenon that we have empirically witnessed during the COVID-19 pandemic, such as slow declines and plateau-like behavior in incidence/prevalence [53, 57, 58]. To achieve the ultimate ambition of developing robust, general theories about human behaviour in relation to infectious diseases [59], we advocate further work in this understudied area to hasten our understanding of the feedback between behaviour, intervention response, policy and the perceived risk from the current epidemiological situation.

Other factors affecting our predictive ability are the inherent uncertainty when fitting models to data, estimates of vaccine delivery, and the unknowable character of future variants. Models will never be a perfect representation of the natural world and when fitting models to data parameter uncertainty will always be present. We represent this parameter uncertainty by showing the 95% prediction interval (which contains 95% of all projections at each time point, ignoring the top and bottom 2.5%). We make use of multiple data sources (reported cases, hospital admissions, hospital occupancy and death) to obtain a more accurate estimation of the parameters, but uncertainties still exist. In addition, given epidemic models often show exponential growth, small uncertainties in this growth rate are magnified by the exponential process generating large upper bounds on the 95% prediction interval - this is especially true when cases are just beginning to rise from a trough as in Roadmap 4a (Fig. 6).

Vaccine uptake and the speed of vaccine delivery also both affect the projected scale of future outbreaks. Inaccuracies in the delivery speed generally impact the short-term dynamics, while inaccuracies in the assumed level of vaccine uptake have longer term consequences due to changing the ultimate level of protection in the population (in a similar manner to errors in estimating vaccine efficacy). While estimates of future vaccine delivery speed were prescribed to the modelling teams, estimates of uptake were instead based on observed historical patterns. However, it is worth noting that while there is highly accurate data on the number of vaccines delivered, the information on the number of people of a given age within England is an estimate, here taken from ONS [50] which is based on the 2011 census. In particular, the recorded number of vaccine first doses given to 55-59 and 60-64 year olds exceeds the estimated population size used in the model (Fig. 2, thin black bar and wide grey bar) and hence requires re-scaling. Given that we are primarily interested in those not vaccinated (and hence more likely to be susceptible to the virus), small errors in the estimation of population size could affect both the model projections and our ability to estimate vaccine efficacy.

Finally, novel variants pose a major problem for robust and accurate prediction - their timing and characteristics cannot be predicted in advance - and hence all the Roadmap documents assume a continuation of the dominant variant.

Such uncertainties (from behaviour, data and variants) highlight our inability to make truly accurate long-term predictions (as opposed to scenario based projections); for example neither the timing of Omicron emergence, the characteristics of this variant in terms of transmission and vaccine protection, nor the public response to this potential health problem could be forecast before the event. This uncertainty highlights why model results should be considered as projections of particular (epidemiologically-plausible) scenarios, rather than predictions or forecasts. To generate true predictions would require models that incorporated the full uncertainty in public behaviour and the emergence of novel variants [60] – leading to prediction intervals that would likely be too large for the results to be useful.

The challenges in producing accurate projections are not unique to England, and are exemplified by the work of the COVID-19 Scenario Modelling Hub in the USA. The COVID-19 Scenario Modelling Hub has specified a set of scenarios to conduct multi-model studies, with these scenarios being designed in consultation with academic modelling teams and government agencies, such as the Centers for Disease Control and Prevention (CDC). Work was convened in multiple rounds with prescribed target outcomes that allowed collective insights to be drawn across the independent model submissions; rounds included an assessment of NPI scenarios over April–September 2021 [61] and resilience to new variants for the period November 2021-March 2022 [62].

While the six Roadmaps had a definite purpose at the time, with hindsight we can consider the lessons learned from this process and how that can inform future modelling. Models were only one part of the advice that fed into the decision-making, and yet was one of the few elements that was publicly available; this level of visibility together with the quantitative nature of the work makes modelling an easy target for those that do not agree with the associated policy decision. Therefore, one of the key issues for the future involves the effective communication of model results, approaches and expectations. Potentially the main message is that our results are a projection of plausible scenarios (not detailed forecasts), and hence we learn more from discrepancies between models and data, than we do from close agreement. When models and data diverge it is usually an indication of inaccuracies in the underlying assumptions, identifying and correcting such errors is how science rapidly progresses, or it may point to unanticipated changes in behaviour, control or transmission.

Associated with the prominence of epidemiological modelling results is the potential for the integration of epidemiological results with other factors (such as social and economic consequences), and for this holistic package to be publicly communicated. A more joined-up approach, balancing all aspects, would provide greater clarity and allow the trade-offs to be more fully explored.

A second key lesson is the huge advantage of multiple groups taking independent approaches, but with a common goal. The Roadmap documents produced by SPI-M-O were a combination of projections from three groups (University of Warwick, the London School of Hygiene and Tropical Medicine, and Imperial College London), this plurality of approaches brings far greater robustness (as highlighted by the Macpherson report [63]) especially when projections are having to be generated under very tight time constraints. Ideally, the involvement of more groups would be beneficial, as was seen with SPI-M-O’s estimates of the reproductive number [64] and Medium Term Projections [4, 65] in the UK and CDC’s COVID-19 Forecasts for the USA [66], but the Roadmaps required longer-term projections which restricted participation to a smaller group of mechanistic models.

It is often quoted that “all models are wrong, but some models are useful” [67], and this is especially true of models used during an unfolding outbreak and attempting to make relatively long-term projections. Therefore it is important to temper our expectations of such models, and realise that departures between model projections and data are also useful policy guides.

Finally, in any epidemic, high-quality and timely data is key. Ideally, anonymised and linked records of testing, cases, vaccination and severe health outcomes would be readily available to those making projections, although this is both ethically and logistically challenging [68, 69]. In general, UK modellers that were part of SPI-M-O had access to a vast amount of information – far larger than in any previous outbreak – although hospital data was generally aggregated, and linkage between data sets only occurred later in the pandemic. Other countries had different approaches to data-sharing [70], but clearly there were many countries where such detailed data were not accessible, although on-line resources became an invaluable tool for monitoring the unfolding epidemic [71].

It is often tempting with hindsight to revisit the decisions made during the pandemic [6, 72], for example we may wish to calculate the effects of taking the relaxation steps faster or slower. While this is possible under the simple assumption that everything else remains fixed, the lesson from trying to forecast precautionary behaviour is that there are multiple inter-related factors. The scale of the ‘pingdemic’ is clearly related to both the social mixing and the number of cases at the time, the slight rise in precautionary behaviour in June 2021 is likely in response to media coverage and concern over the Delta variant. The desire to understand the implications of different policy decisions at different times therefore necessitates a larger more holistic view of the epidemic where the broader consequences of any policy change are reflected in projections.

These Roadmap projections [14–19], together with the projections from other groups [20–25], have provided quantitative support for the steady relaxation of mitigation measures. In general, Roadmap modelling suggested that hospital admissions would not rise to the extreme levels observed during the first and second waves (with peak admissions of 3099 and 4134 in England respectively) following the relaxation steps. The exception to this was Roadmap 4a [17, 23], where uncertainty over the Delta variant (especially the protection afforded by vaccination) led to a range of projections many with high peak hospital admissions (Fig. 6). As such, these model projections have proved to be a useful policy tool, providing a mechanism for translating epidemiological knowledge and uncertainties into medium-term assessments of the likely range of public health burden.

## Methods

Although the development of the Warwick SARS-CoV-2 transmission and COVID-19 disease model has been described elsewhere in extensive detail [5, 7, 13, 28], here we summarise the main salient components and the method of parameter inference.

### Model overview

The model is built around the traditional deterministic SEIR (Susceptible, Exposed, Infectious, Recovered) model framework [29], with three exposed classes to capture the distribution of times from infection to becoming infectious [30], and splitting the infectious group into symptomatic and asymptomatic infection (Fig. 9a). To this simple model we add additional structure to capture the effects of restricted social interaction whilst maintaining household transmission [5], and this fundamental model is then ‘replicated’ twenty-one times to mimic five-year age-groups (0 *−* 4, 5 *−* 9, …, 100+). The model is written as a large number of ODEs (ordinary differential equations).

**Fig. 9:**
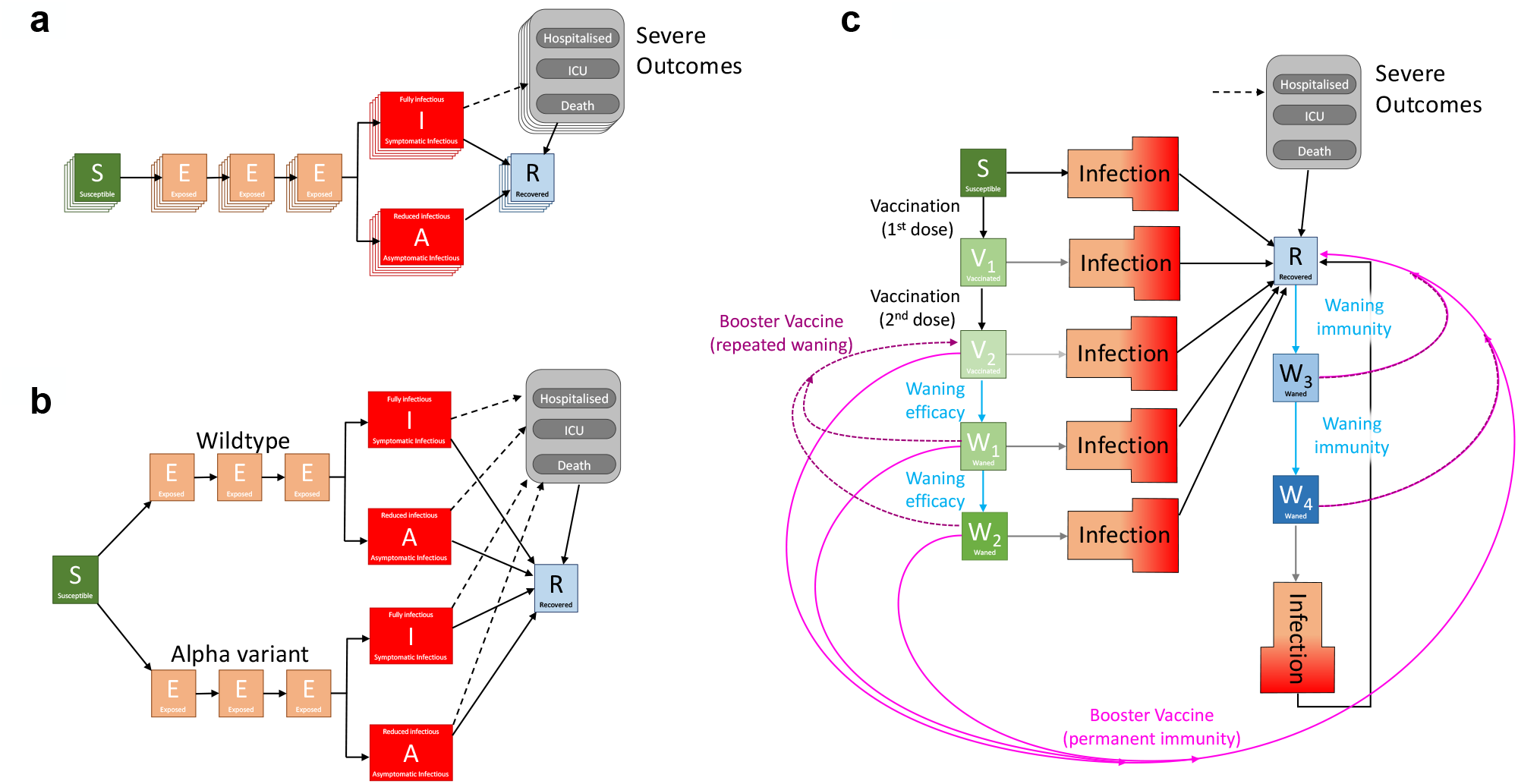
Caricatures of the development of the Warwick SARS-CoV-2 transmission and COVID-19 disease model. **a** The basic *SEIR*-type structure of the model, with three exposed classes, infectious individuals partitioned into symptomatic (*I*) and asymptomatic (*A*), and with severe outcomes driven by symptomatic cases; the multiple layers represent age-structure. **b** The inclusion of an extra variant increases the number of infection classes that are required (here, the layers representing age-structure have been removed for clarity). **c** The introduction of vaccination, waning immunity (blue) and booster vaccination (pink) dramatically increases the complexity of the model, hence we group all infected classes for clarity and do not show the multiple variants or age-structure).

This basic model was sufficient for the early waves of infection (from January to November 2020) with a single variant and without vaccination. During this early phase of the pandemic, the main driving parameter was the level of precautionary behaviour, which determined the level of social-mixing and therefore the scale of transmission outside the household [13], although we also fitted a number of other parameters (including case:hospitalisation and case:mortality ratios, age-dependent effects and the relative strength of asymptomatic compared to symptomatic transmission). From the age-structured symptomatic class, we calculate the number of severe outcomes (hospital admissions, intensive care unit admissions and deaths), which are both key public health observables and measures of concern for this pandemic, although these quantities do not impact the transmission dynamics (Fig. 9a). The fitting is performed in a Bayesian framework, matching the data on the daily hospital admissions, hospital occupancy, ICU occupancy, deaths and proportion of community (Pillar 2) tests that are positive in each of the seven National Health Service (NHS) regions of England to a Poisson distribution with a mean given by the ODE model.

From late 2020, variants and vaccination increased the dimensionality of this model. Each new variant required a duplicate of all the infected model classes (Fig. 9b, shows the addition of the Alpha variant), to capture differences in transmission and risks of severe outcomes; the rise of each variant was captured by additionally fitting to the proportion of S-gene target failures (a measure of variant-type) from TaqPath PCR testing [31, 32]. Early Roadmap models (1-3) just modelled wildtype and Alpha variants, while Roadmaps 4a, 4b and 5 also included the Delta variant. The start of the vaccination campaign in December 2020 necessitated a further partitioning of the population by vaccination status (first three elements in Fig. 9c), allowing us to capture both the reduced risk of infection and the reduced risk of severe outcomes. For the final Roadmap, waning levels of protection both in terms of vaccine-induced and infection-induced immunity were added (generating additional elements in Fig. 9c, with waning immunity processes shown as blue arrows) and booster vaccination (shown as pink arrows in Fig. 9c) leading to long-lasting immunity (bright pink solid lines) or repeated waning (dark pink dotted lines).

Each of the Roadmaps required assumptions to be made about how changes in policy associated with Steps 1-4 would translate into changes in precautionary behaviour. For early Steps (and hence early Roadmaps), the precautionary behaviour after each Step was estimated with reference to historical values, matching future restrictions with comparable restrictions in the past and assuming that population adherence would be similar. For later Steps, it was assumed that either a moderate amount of precautionary behaviour would remain after Step 4 (the assumption applied in Roadmaps 1-4a, and sensitivity to this level was generally assessed) or that behaviour would gradually return to prepandemic levels over several months (Roadmaps 4b and 5). In the descriptions that follow, we quote the estimates associated with each Step in terms of R excluding immunity (*R*_*ei*_), which provides an unambiguous measure of transmission during each period. This is inversely related to the precautionary behaviour (high precautionary behaviour leads to low *R*_*ei*_ and lower transmission) but the relationship is non-linear being derived from an eigenvalue of transmission matrices; with precautionary behaviour providing a proportional scaling between transmission matrices associated with pre-pandemic and stringent lockdown behaviour.

One of the key sets of parameters feeding into each Roadmap model was the protective effects of vaccination. This is partitioned into five elements: protection against infection; protection against symptomatic disease; protection against requiring hospital admission; protection against death; and the reduction in onward transmission for vaccinated individuals who do become infected (Table 1). For Roadmaps 1 to 3 only protection against the Alpha variant is required; whereas in Roadmaps 4a-5 projection against the Delta variant is also included. For Roadmap 5, we also model waning immunity (using 3 different assumptions) and boosting (2 assumptions).

We now consider the models used in each Roadmap in more detail, contrasting the changes that were needed to keep pace with our knowledge of the pandemic.

### Roadmap 1, 6th February 2021

During early modelling, upon which Roadmap 1 was based, data on the degree of vaccine protection was extremely limited. Clinical trials had been against the wild-type virus, whereas at this time the population in England was predominantly infected with the Alpha variant (Table 1). We used two levels of vaccine protection, which were independent of the type of vaccine (AstraZeneca or Pfizer) administered, and due to lack of evidence, the modelled vaccine provided no additional protection against hospital admission or death beyond blocking infection (although [14] appendix 13 did consider a scenario of very high efficacy against severe disease). The central assumptions were chosen to be close to the mid-points of the confidence intervals for efficacy values estimated at the time [33–35]. The cautious assumptions were chosen as half this protective effect for infection blocking (where there was limited data) and closer to the lower confidence intervals for efficacy against symptomatic disease. The central and cautious assumptions also differ in the assumed level of vaccine uptake in 18-49 year olds, assuming 85% and 75%, respectively; whereas in all projections vaccine uptake was modelled as 95% in the over 80s, and 85% in those aged 50-79.

Four timing scenarios were investigated that set the scene for the relaxation of controls, each based on 5 Steps (although we note that the adopted policy on 22nd February 2021 was for 4 Steps [12]) – from a return of children in primary and secondary schools to full-time education (Step 1) to an absence of all controls (Step 5). Only the three timing scenarios (scenarios Two, Three and Four) that most closely correspond to the realised relaxation are shown here. For each relaxation step, there was the challenge of determining the level of social mixing and hence the modelled degree of transmission. Stage 1 was based upon the January lockdown data (but with children returning to schools), while Stage 4 was based upon behaviour in September 2020; Stages 2 and 3 were a 70:30 and 30:70 mix of Stages 1 and 4 respectively. Stage 5, which removed all controls was considered at three levels of transmission (low, medium and high) - which we achieved by setting precautionary behaviour at 20%, 10% or 0%. These levels of precautionary behaviour (Fig. 1a) are better characterised in terms of the (model independent) reproductive ratio excluding immunity *R*_*ei*_: low (*R*_*ei*_ = 3.0), medium (*R*_*ei*_ = 3.6), and high (*R*_*ei*_ = 4.0). Here, the reproductive ratio excluding immunity corresponds to the average number of secondary cases produced by an average infectious individual when the entire population is susceptible, but when some degree of control potential exists; as such it provides a useful model-independent quantification for the strength of controls - and later for the impact of new variants.

### Roadmap 2, 29th March 2021

By the time the second Roadmap document was produced, the relaxation pathway taking England from lockdown to the complete absence of controls was clearly defined, with four Steps between the January 2021 lockdown and removal of all social mixing restrictions. The second Roadmap document considered a number of sensitivity analyses, including the impact of seasonal forcing (which we present in this paper, as including a moderate level of seasonality became a default assumption in future work). Estimates of social mixing within each of the relaxation steps was still challenging, having to be based on assumptions about how the population was expected to react to each of the relaxation step changes. From consideration of previous controls and the inferred levels of precautionary behaviour, we assumed that each Step would lead to a discrete change in precautionary behaviour (75% of the early March 2021 level in Step 2, 35% in Step 3 and 15% in Step 4), which we again translate into *R* excluding immunity: Step 1 *R*_*ei*_ = 1.49 (CI 1.27 *−* 1.65); Step 2 *R*_*ei*_ = 1.76 (CI 1.59 *−* 1.90); Step 3 *R*_*ei*_ = 2.53 (CI 2.40 *−* 2.69); and Step 4 *R*_*ei*_ = 2.97 (CI 2.79 *−* 3.14). We note that the assumptions for Step 4 (complete absence of legislation) is most comparable to the medium mixing assumption during Stage 5 in Roadmap 1.

By the end of March 2021, the parameters for vaccine efficacy (against Alpha) had also been refined, with new estimates for the protection against severe disease (hospital admission and death) and the differences between the AstraZeneca and Pfizer vaccines (Table 1) based on early reports from Israel and England [36–39]. The reduction in onward transmission (from individuals that have been vaccinated but then later infected) was still unknown due to the difficulties of estimating this quantity; taking a precautionary approach this reduction was set to zero (Table 1). The absence of attenuation in onward transmission from infectious but vaccinated individuals reduces the estimated impact of vaccination in terms of indirect protection, although there is still considerable indirect protection from both reduced levels of infection and lower levels of symptomatic disease.

### Roadmap 3, 4th May 2021

By late April 2021, the clear exponential decline in cases and severe disease was well established. Vaccine efficacy for the Alpha variant was also reasonably well defined (Table 1), although higher and lower efficacies were considered as a sensitivity analysis. For this Roadmap, modelling the easing of restrictions for England [16], two major refinements were made to the model structure: the inclusion of a reduction in onward transmission from vaccinated individuals who become infected (although the precise scale of this reduction was, and remains, difficult to estimate); and the inclusion of 10% seasonality as a default - reducing the scale of any summer outbreaks.

The model still assumed that each Step would lead to a discrete change in the level of precautionary behaviour estimated at 55% of the value at the end of the third lockdown in early March 2021 for Step 3 and 15% for Step 4. This can be translated into *R* excluding immunity: Step 1 *R*_*ei*_ = 1.66 (CI 1.42 *−* 1.83); Step 2 *R*_*ei*_ = 1.88 (CI 1.64 *−* 2.10); Step 3 *R*_*ei*_ = 2.41 (CI 2.25 *−* 2.57); and Step 4 *R*_*ei*_ = 3.51 (CI 3.31 *−* 3.71); - with Steps 1 and 2 informed by the most recent data.

### Roadmap 4a, 8th June 2021

We produced our initial Roadmap modelling for the Step 4 easing restrictions in England using data up to 4th June 2021 [17], less than a month after Delta was declared a Variant of Concern (6th May 2021 [41]) and around the time that Delta became the dominant variant in the UK. These circumstances inherently meant that the projections were being made with relatively little data on the transmission advantage of Delta over Alpha and with extremely limited data on the protection offered by vaccination. We estimated that the Delta variant had a 56% (CI 34-81%) transmission advantage over the Alpha variant, which produced a good match to the early increase in S-gene positive cases – a rapid signal from PCR testing of the spread of Delta [31]. It was thought at this time that a single dose (of either vaccine) would offer far less protection against Delta compared to Alpha. The lower protection afforded by the vaccine towards the Delta variant was reflected in the model assumptions; the Central vaccine assumptions were taken from the preliminary PHE estimates [43], where a single dose of vaccine only generated 34% protection against Delta, compared to 60% against Alpha (Table 1). A range of vaccine efficacy parameters were explored to capture the uncertainty in these parameters.

In this Roadmap we also still assumed that Step 4 (and the removal of all legal restrictions on behaviour) would lead to a large increase in social mixing, and hence a large increase in transmission. The assumed mixing for Step 3 and 4 would lead to R excluding immunity estimates for the more transmissible Delta variant of *R*_*ei*_ = 3.85 (CI 3.63 *−* 4.10) and *R*_*ei*_ = 5.66 (CI 5.4 *−* 5.95) respectively. Although Step 3 had begun on 17th May 2021, there was still insufficient evidence from cases and more severe disease to infer the level of precautionary behaviour.

This uncertainty is acknowledged in the Roadmap document [17] where we state that any delay to the relaxation roadmap would have three main epidemiological advantages: (a) delay and reduce the epidemic peak providing more time for additional vaccination and protection; (b) provide additional time to understand the degree to which the vaccine protects against the Delta variant; (c) allow more confidence to separate the impacts of the Delta variant from changes due to Step 3. There was also concern at the time that Delta had a slightly elevated case hospitalisation rate when compared with Alpha [42], which would increase associated projections by a multiplicative scaling.

### Roadmap 4b, 6th July 2021

Our second Roadmap modelling focusing on easing restrictions for England (Step 4) used data up to 2nd July 2021 [18], by which point far more information had been gathered on vaccine efficacy [44], suggesting that protection against hospitalisation was closer to the most optimistic of earlier assumptions, whilst protection against infection from the Pfizer vaccine was again very high (Table 1). Despite the availability of an additional month of data to refine vaccine efficacy estimates, within this Roadmap we still considered more cautious and more optimistic assumptions as part of the sensitivity analysis given the importance of vaccine efficacy parameters. The extra data (since Roadmap 4a) also allowed better inference of the precautionary behaviour in Step 3 and hence a refinement of the transmission advantage of Delta over Alpha, with an updated estimate of 68% (38-86%).

For the first time in our Roadmap modelling we also had sufficient data to establish that behaviour did not respond rapidly to legislation and consequently that individuals were unlikely to return to pre-COVID mixing as soon as restrictions were lifted. However, this gave rise to substantial uncertainty about the future, and as such, multiple pathways to a complete return to normality were investigated, ranging in both the initial drop once Step 4 occurred and the time taken to reach pre-COVID mixing.

### Roadmap 5, 12th October 2021

In our final Roadmap document of 2021 [19], the aim was to consider the likely dynamics for Autumn and Winter 2021. Echoing the previous Roadmap document [18], we considered the return to pre-COVID mixing over multiple time-scales (from December 2021 to June 2022).

The largest change in model structure for this Roadmap was the inclusion of waning vaccine efficacy - supported by growing evidence about the decline in protection over time [45–47]. To combat this decline in immunity, booster vaccines were modelled in this Roadmap document as being offered to all those over 50 years old at a rate of 1.3 million doses per week. Therefore, while the early protection against the Delta variant offered by vaccination was by this point well-defined (Table 1), we now encountered a new set of uncertainties about waning and boosters. Firstly, would vaccine efficacy from the first two doses continue to decline to zero, or would protection asymptote to a low (but non-zero) level (Table 1)? To address this question we made three different assumptions about the long-term level of vaccine protection which corresponded to three different half-lives for the rate of waning (180, 270 and 460 days respectively) to match the available data [45–47]. Secondly, there was also considerable uncertainty about the strength and duration of protection after the booster. We modelled two limiting scenarios (Table 1): in the first the booster resets vaccine protection to the level after the second dose (nullifying the waning effect after the primary vaccine doses), but efficacy then wanes as before; in the second the action of boosters provided high levels of long-lasting protection [48].

### Similarities and differences between Roadmap models

In summary, Roadmap models 1-3 (generated on 6th February, 29th March and 4th May 2021) were remarkably similar in structure, accounting for wildtype and Alpha variants together with the ongoing rollout of vaccination. The main improvements between the models were the increasingly reliable estimates of vaccine protection, and the historic behaviour at previous steps on which to base projections. Roadmap 4 (8th June 2021) necessitated the inclusion of the Delta variant (expanding the model complexity and the number of vaccine parameters required). Roadmap 4b (6th July 2021) had more accurate estimates of vaccine protection against Delta, and for the first time modelled a slow public response to each step potentially lasting many months. Finally, for Roadmap 5 (12th October 2021) we included both waning immunity and booster vaccines, which again increased the underlying dimensionality of the model.

### Data Availability

Data on cases were obtained from the COVID-19 Hospitalisation in England Surveillance System (CHESS) data set that collects detailed data on patients infected with COVID-19. Data on COVID- 19 deaths were obtained from Public Health England. These raw data contain confidential information, with public data deposition non-permissible for privacy reasons. The CHESS data resides with the National Health Service (www.nhs.gov.uk) whilst the death data are available from Public Health England (www.phe.gov.uk). Again these raw data contain confidential information, with public data deposition non-permissible for privacy reasons. The ethics of the use of these data for these purposes was agreed by Public Health England with the Governments SPI-M-O / SAGE committees. Processed data (which is more aggregated) is freely available from the UK Coronavirus dashboard: https://coronavirus.data.gov.uk/

## Data Availability

Data on cases were obtained from the COVID-19 Hospitalisation in England Surveillance System (CHESS) data set that collects detailed data on patients infected with COVID-19. Data on COVID-19 deaths were obtained from Public Health England. These data contain confidential information, with public data deposition non-permissible for socioeconomic reasons. The CHESS data resides with the National Health Service (www.nhs.gov.uk) whilst the death data are available from Public Health England (www.phe.gov.uk). The ethics of the use of these data for these purposes was agreed by Public Health England with the Governments SPI-M(O) / SAGE committees. More aggregate data is freely available from the UK Coronavirus dashboard: https://coronavirus.data.gov.uk/

## Code Availability

Examples of the code needed to generate Roadmaps 1-4b without waning and boosters (as shown in Fig. 9b), and to generate the more complex Roadmap 5 (as shown in Fig. 9c) can be found at https://github.com/MattKeeling/RelaxationRoadmaps.git [73]

## Ethical Considerations

Data from the CHESS and SARI databases were supplied after anonymisation under strict data protection protocols agreed between the University of Warwick and Public Health England. The ethics of the use of these data for these purposes was agreed by Public Health England with the Government’s SPI-M-O / SAGE committees.

## Acknowledgements

We would like to thank many people for their thoughts, comments and help during the lengthy process of producing the six Roadmap documents. Members of the JUNIPER consortium have offered continual support, encouragement and advice throughout this period; Graham Medley and Angela McLean as chairs of SPI-M-O provided invaluable input into the development of these Roadmaps; the SPI-M-O secretariat for their continued support (Paul Allen, Tom Irving, Libby Richards, Mat Katz, Jen Huynh and Alastair Ikin); members of PHE/UKHSA that have assisted with data access (Andre Charlotte and Nick Gent) and estimates of vaccine efficacy (Nick Andrews, Jamie Lopez Bernal and Mary Ramsay); and members of DSTL who have helped with data processing (Ronni Bowman and Phillippa Spencer).

MJK, LD and MJT were supported through the JUNIPER modelling consortium [grant number MR/V038613/1]; MJK and SM were supported by the National Institute for Health Research (NIHR) [Policy Research Programme, Mathematical and Economic Modelling for Vaccination and Immunisation Evaluation, and Emergency Response; NIHR200411]; MJK, LD, MJT and EMH were supported by the Medical Research Council through the COVID-19 Rapid Response Rolling Call [grant number MR/V009761/1] MJK is affiliated to the National Institute for Health Research Health Protection Research Unit (NIHR HPRU) in Gastrointestinal Infections at University of Liverpool in partnership with UK Health Security Agency (UKHSA), in collaboration with University of Warwick. MJK is also affiliated to the National Institute for Health Research Health Protection Research Unit (NIHR HPRU) in Genomics and Enabling Data at University of Warwick in partnership with UK Health Security Agency (UKHSA). The views expressed are those of the author(s) and not necessarily those of the NHS, the NIHR, the Department of Health and Social Care or UK Health Security Agency.

## Author Contributions

M.J.K: Conceptualisation, Data curation, Formal analysis, Funding acquisition, Methodology, Software, Validation, Visualisation, Writing—Original draft, Writing—Review and Editing. L.D.: Conceptualisation, Data curation, Funding acquisition, Methodology, Software, Validation, Writing—Review and Editing. M.J.T.: Conceptualisation, Funding acquisition, Writing— Review and Editing. E.H.: Conceptualisation, Data curation, Methodology, Software, Validation, Writing—Review and Editing.

S.M. Conceptualisation, Data curation, Formal analysis, Methodology, Software, Validation, Visualisation, Writing—Review and Editing.

